# NONENDOSCOPIC DETECTION OF BARRETT’S ESOPHAGUS IN PATIENTS WITHOUT GERD SYMPTOMS

**DOI:** 10.1101/2025.03.26.25324651

**Authors:** Amitabh Chak, Komal Keerthy, Gi-Ming Wang, Wendy Brock, Beth Bednarchik, Rajesh Guptha, Suman Verma, Helen Moinova, Curtis Tatsuoka, John Dumot, Sapna Thomas, Joseph E. Willis, Sanford Markowitz

## Abstract

**Background:** Screening efforts for Barrett’s Esophagus (BE) predominantly focus on performing upper endoscopy (EGD) on patients with gastroesophageal reflux disease (GERD) symptoms who have additional risk factors for BE. However, cost and invasiveness preclude EGD in those who have no prior GERD symptoms, despite having other risk factors, representing missed opportunities for BE screening in individuals who account for approximately 40% of the patients who eventually develop esophageal adenocarcinoma (EAC).

**Aim:** The aim of this study was to evaluate if non-endoscopic methods can enable BE detection in an at-risk population without GERD symptoms.

**Methods:** Patients presenting for colonoscopy who had not undergone previous EGD plus had ≥3 BE risk factors (from among age ≥50 years, male sex, white race, smoking history, family history of BE/EAC, or central obesity) without chronic GERD symptoms were prospectively recruited for non-endoscopic screening. Trained nurses administered the EsoCheck (Lucid Diagnostics) encapsulated balloon. Samples were assayed with the EsoGuard BE detection methylated DNA marker panel (Lucid Diagnostics). Patients with a positive result were offered standard-of-care EGD, while patients with a negative EG result were offered free of cost research EGD. Positive predictive value (PPV), negative predictive value (NPV), and BE prevalence were calculated.

**Results:** The mean age of the 132 study subjects was 60.7 years, 129 (98%) were white, 124 (94%) were male, 71 (54%) had a prior smoking history, 46 (35%) were centrally obese, and 5 (4%) reported a family history. EsoCheck was successfully administered in 124 (94%) and the EsoGuard methylated DNA marker panel could be assayed in 120 (97%) of the samples. Thirty-four assays were positive of which 27 underwent a follow-up EGD and BE was identified in 9, PPV = 33% [17%, 54%] subjects. EGD was also performed in 22 of the 86 subjects whose assays were negative and none of them had BE, NPV = 100% [85%, 100%]. A logistic regression model fitted to impute the presence of BE estimated the PPV as 27% [13%, 44%], NPV as 98% [92%, 100%], and BE prevalence as 8.4% [4.5%, 14.3%].

**Conclusion:** Patients without chronic GERD who have ≥3 BE risk factors have a moderately high prevalence of BE. Non-endoscopic detection can effectively identify BE, enabling expansion of screening to this larger at-risk population. Those with a negative EG assay have a low likelihood of BE.

---

Barrett’s esophagus (BE) is the only known precursor of esophageal adenocarcinoma (EAC) with modeling studies suggesting that all EACs arise from this precursor.^1^ Thus, guidelines traditionally recommend sedated upper endoscopy (EGD) in patients with chronic gastroesophageal reflux disease (GERD) symptoms and additional risk factors for BE as a screening strategy for prevention and/or early detection of EAC.^2-4^ The limitations of these guidelines, which have been in place for decades, are clear, as most BE remains undetected prior to the diagnosis of EAC and the prognosis of this cancer remains poor. A strategy based on only screening those with chronic GERD symptoms will also have only limited impact, as 40% of EACs occur in patients who don’t report GERD.^5,6^

Our team successfully developed a selective distal esophageal sampling device, now commercially available as EsoCheck® (EC, from Lucid Diagnostics, New York, NY) combined with a methylated DNA biomarker panel, commercialized as EsoGuard® (EG, LucidDx Labs, Lake Forest, CA) that enables non-endoscopic detection of BE.^7^ This combination detected early non-dysplastic BE (NDBE) with 90% sensitivity and 92% specificity.^7^ A follow-up validation study demonstrated 85% sensitivity and 85% specificity for EsoCheck/EsoGuard across BE of all stages and lengths.^8^ This office-based detection technology is already being clinically used to expand access to BE screening among patients with chronic GERD symptoms meeting guideline criteria.^2^ In fact, a study of veterans with chronic GERD symptoms who met screening criteria found that EsoCheck/EsoGuard had a PPV and NPV of 33% and 99%, respectively.^9^ To determine if non-endoscopic methods could further enhance screening in the full population at-risk for BE, we decided to non-endoscopically screen patients who had multiple risk factors for BE but did not report chronic GERD symptoms.

## METHODS

The study methodology was approved by the Institutional Review Board for Human Investigation at University Hospitals Cleveland Medical Center and was registered at clinicaltrials.gov as NCT04880044. The electronic medical record was searched for patients who were scheduled for outpatient colonoscopy to identify eligible patients as: (1) those without chronic GERD (i.e., had no diagnosis of GERD and/or had not been on acid suppressive medications for ≥ 5 years); (2) had three or more of the following BE risk factors: age ≥ 50 years, male gender, white race, smoking history, obesity (BMI ≥ 30), or family history of BE or EAC; and (3) had not undergone previous EGD. Patients on anticoagulant medications, history of head and neck cancer, and suspected esophageal varices were excluded. Eligible patients were approached for study participation by a research coordinator. Those who consented were requested to fill out a brief survey confirming they had not had regular heartburn or regurgitation for ≥ 5 years. Subjects also confirmed the ability to swallow pills and lack of dysphagia. EsoCheck distal esophageal cell sampling was performed by the research coordinator according to the device instructions for use and the sample was sent for analysis with the EsoGuard assay. If the EsoGuard assay result came back positive, the patient was informed of the result and recommended an EGD. Patients with negative results were told to discuss with their primary physicians whether they should undergo an EGD and were offered a free-of-cost study EGD with biopsy of the gastroesophageal junction. For those who underwent EGD the presence of suspected BE defined as ≥ 1 cm of columnar mucosa in the tubular esophagus was noted and confirmed by the presence of intestinal metaplasia on biopsies obtained from the tubular esophagus. Biopsies were also obtained from the squamocolumnar junction when there was no visible columnar mucosa in the tubular esophagus. The length of BE was classified using Prague criteria. A blinded adjudicator reviewed endoscopic images of all patients classified as having an irregular Z-line and short segment C0M1 or C0M2 BE. The adjudicators interpretation of length was considered final. The presence of intestinal metaplasia in biopsies obtained from a normal or irregular Z-line was classified as cardia intestinal metaplasia (CIM).

All subjects were administered a modified Mayo GERQ to confirm the absence of chronic heartburn or reflux defined either as (a) no heartburn or regurgitation, or (b) less than 5 years of symptoms or less than weekly symptoms if symptoms were reported.^10^ The modified questionnaire also asked patients if they smoked or had previously smoked, had a family history of BE or EAC, and to report pant waist size in inches. Age, gender, race, and body mass index (BMI) were collected from the electronic medical record. Obesity was defined as waist size of 40 inches or more in men, 35 inches or more in women, and/or BMI ≥ 30 kg/m^2^.

An analytic logistic regression model was built to classify the unknown phenotype of patients who did not undergo an EGD. The training set for the model consisted of data from the subset of patients who underwent EGD and had confirmed phenotypes of BE or non-BE. First, missing values, such as smoking history, family history and waist measurement, were imputed using predictive mean matching (PMM) method. The EsoGuard outcome was withheld from imputation, as it could be excessively aggressive, considering our aim is to create a link between EsoGuard and presence of BE in the initial analysis. The EsoGuard outcome was also excluded from the logistical model, as the presence of BE was determined exclusively by patient-related factors; however, it was employed for imputing these patient factors. Second, the model’s performance was validated by an area under the ROC curve (AUC=0.747) and Brier Score (0.128) to determine the model’s ability to discriminate between BE and normal non-BE phenotypes and to make probabilistic predictions. Finally, after setting a threshold of classification to 0.4 for presence of BE, determined by the ROC curve, the unknown phenotypes were predicted using our fitted model.

## RESULTS

The study was conducted between January 2022 to February 2025. Of 258 eligible subjects without chronic GERD who had 3 or more risk factors for BE, 132 (51%) subjects agreed to undergo non-endoscopic screening. The majority were men (94%), white (98%), and mean age was 60.7 years (Table 1). EsoCheck was swallowed and the distal esophagus was sampled in 124 (94%) subjects. The EsoGuard assay was positive in 34 (27%), negative in 86 (69%), and could not be determined in 4 (3%) subjects due to insufficient sample. Subjects with a positive EsoGuard assay were significantly older, were more likely to report a positive family history, and tended to have a history of smoking (Table 1).

**Table 1:** Demographics and Risk Factors in Study Subjects.

| **Characteristic** | **Overall**<br>**N = 132<sup>1</sup>** | **EsoGuard**<br>**Negative**<br>**N = 86<sup>1</sup>** | **EsoGuard**<br>**Positive**<br>**N = 34<sup>1</sup>** | **EsoGuard**<br>**Unknown**<br>**N = 12<sup>1</sup>** | **p-value<sup>2</sup>** |
| --- | --- | --- | --- | --- | --- |
| **Sex** |  |  |  |  | 0.729 |
| Male | 124 (94%) | 81 (94%) | 31 (91%) | 12 (100%) |  |
| Female | 8 (6.1%) | 5 (5.8%) | 3 (8.8%) | 0 (0%) |  |
| **Mean Age (SD)** | 60.7 (7.5) | 59.5 (7.7) | 64.5 (6.6) | 59.4 (4.6) | 0.001 |
| **Race** |  |  |  |  | 0.727 |
| White | 129 (98%) | 84 (98%) | 33 (97%) | 12 (100%) |  |
| Black or African American | 2 (1.5%) | 1 (1.2%) | 1 (2.9%) | 0 (0%) |  |
| Hispanic | 1 (0.8%) | 1 (1.2%) | 0 (0%) | 0 (0%) |  |
| **Smoking history** |  |  |  |  | 0.095 |
| No | 60 (45%) | 40 (47%) | 13 (38%) | 7 (58%) |  |
| Yes | 71 (54%) | 46 (53%) | 21 (62%) | 4 (33%) |  |
| Unknown | 1 (0.8%) | 0 (0%) | 0 (0%) | 1 (8.3%) |  |
| **Family history** |  |  |  |  | 0.030 |
| No | 120 (91%) | 81 (94%) | 30 (88%) | 9 (75%) |  |
| Yes | 5 (3.8%) | 2 (2.3%) | 3 (8.8%) | 0 (0%) |  |
| Unknown | 7 (5.3%) | 3 (3.5%) | 1 (2.9%) | 3 (25%) |  |
| **Obesity** |  |  |  |  | 0.166 |
| No | 85 (64%) | 58 (67%) | 21 (62%) | 6 (50%) |  |
| Yes | 46 (35%) | 28 (33%) | 13 (38%) | 5 (42%) |  |
| Unknown | 1 (0.8%) | 0 (0%) | 0 (0%) | 1 (8.3%) |  |
| **BE** |  |  |  |  | <0.001 |
| No | 40 (30%) | 22 (26%) | 18 (53%) | 0 (0%) |  |
| Yes | 9 (6.8%) | 0 (0%) | 9 (26%) | 0 (0%) |  |
| Unknown | 83 (63%) | 64 (74%) | 7 (21%) | 12 (100%) |  |

<sup>1</sup>n (%); Mean (SD)
<sup>2</sup>Fisher's exact test; Kruskal-Wallis rank sum test

**Table 2:** PPV and NPV of EsoGuard for predicting presence of BE at endoscopy.

| **Approach** | **PPV [95% C.I.]** | **NPV [95% C.I.]** | **Prevalence [95% C.I.]** |
| --- | --- | --- | --- |
| **Restricted to subjects who had EGD** | 0.333<br>[0.165, 0.540] | 1.000<br>[0.846, 1.000] |  |
| **Imputed values from entire study cohort** | 0.265<br>[0.129, 0.444] | 0.976<br>[0.918, 0.997] | 8.40%<br>[4.27%, 14.53%] |

Twenty seven (79%) of 34 EsoGuard positive subjects underwent EGD and BE was diagnosed in 9. Using Prague criteria, the BE was assessed to be C0M1 in 6, C0M2 in 2, and C5M7 in 1 subject.^11^ Of the 86 EsoGuard negative subjects, 22 (26%) elected to undergo a free of cost EGD and none of them had BE. CIM was present in 3 of the 18 (17%) EsoGuard positive subjects without BE and in 4 of the 22 (18%) EsoGuard negative subject patients. Erosive esophagitis was noted in 6 (22%) EsoGuard positive and 2 (9%) EsoGuard negative subjects (p = 0.40). The positive predictive value (PPV) of EsoGuard in those who underwent EGD was 33% (95% CI, 17% - 54%) and negative predictive value was 100% (95% CI, 85% - 100%). In these non-GERD patients with 3 or more risk factors, the logistic regression model provided overall imputed PPV as 27% (95% CI, 13% - 44%), NPV as 98% (95% CI, 92% - 100%), and BE prevalence as 8.4% (95% CI, 4.3% - 14.5%).

## DISCUSSION

The development and availability of non-endoscopic office-based testing are poised to greatly enhance BE screening, enabling more widespread prevention of EAC. This study evaluated a viable approach to address a group of at-risk subjects who have previously not undergone BE screening and are unlikely to undergo EGD due to absence of reflux symptoms. The non-endoscopic BE detection test had a PPV of 33% and NPV of 100% in patients without GERD who had three or more risk factors for BE, remarkably similar to the PPV of 33% and NPV of 99% measured in a VA study of patients with GERD^9^ who met traditional guideline criteria for screening.^2^

Recommendations for BE screening have traditionally been focused on adults with symptomatic chronic GERD, which is a strong risk factor for BE and EAC and often brings patients to clinical attention. Guidelines recommend EGD in those with chronic GERD who have additional risk factors for BE or EAC, which include age >50 years, central obesity, smoking, white race, male sex, and family history.^2-4^ However, patients without chronic GERD symptoms who have three, four, or more other BE risk factors may have similar odds for developing BE.^2,12,13^ Indeed, the 2022 AGA Practice Update newly recommends BE screening for individuals with any 3 or more risk factors, and our study now provides the first opportunity for rigorous testing of a practical, non-endoscopic screening approach in this risk group.^14^

Although a strategy of screening patients regardless of the presence of chronic GERD symptoms may not be as cost efficient, it is clinically more effective, because by additionally capturing the 40% of patients who develop EAC without preceding GERD, our proposed non-endoscopic approach reaches the nearly 100% of the US population at risk for developing EAC. To this end, analysis of risk factors from an NCI sponsored multi-institutional study demonstrated that EAC patients with (n=313) or without GERD (n=163) show the same age distribution and same risk profiles of: male gender, white race, smoking, obesity, family history of BE/EAC.^15^ Further, over 80% of EAC patients without GERD had three or more risk factors, with 34% having only three. A recent study that modeled the cost burden of BE screening determined that a strategy based on screening all adults with GERD plus two additional risk factors affects ~23 million, or 10% of U.S. adults; whereas, a strategy of screening adults between the ages of 50-75 with two additional risk factors, similar to the population screened in this study, impacts ~ 66 million, or 29% of U.S. adults.^16^

An intriguing secondary finding in our study was the proportion of both EG positive and negative patients who had erosive esophagitis despite not having chronic “symptomatic” GERD. One limitation of our study is that patients undergoing colonoscopy who elect to undergo research screening may be more motivated and may not represent the general population or perhaps elected to undergo screening because they subconsciously had GERD symptoms that they did not acknowledge. It is also possible that a fair proportion of adults experience GERD on a regular basis but have become so habituated that they are not conscious of their symptoms. Perhaps patients who are said to develop BE or EAC despite not being “symptomatic” for GERD should be better classified as patients who are not conscious of their GERD symptoms.

Although CIM is likely a precursor of adenocarcinoma of the gastroesophageal junction, screening for CIM and subsequent surveillance is not recommended because the risk of progression is quite low and the cost of endoscopic surveillance would be inordinately high. Risk factors for junctional adenocarcinoma are similar to those of adenocarcinoma of the esophagus.^17^ Our study found that CIM was present in nearly one fifth of subjects who were EsoGuard positive but also in subjects who were EsoGuard negative. If non-endoscopic molecular risk markers that predicted which patients with CIM are at risk of progression to high grade dysplasia or cancer could be developed, then non-endoscopic identification of CIM in an at risk population could be used to develop clinical strategies for prevention and early detection of junctional cancers.

Another limitation of our study was that the endoscopist performing the EGD was aware of the results of the EsoGuard assay. This could have potentially resulted in a bias in interpreting endoscopic findings as representing C0M1 BE versus an irregular Z line. We attempted to minimize this bias by retrospective blinded review of the stored images at endoscopy. Although the blinded review did not change any diagnosis, it is still possible that some CIM were misclassified as C0M1 BE and vice versa.

Our study demonstrates that non-endoscopic screening can be used to perform selective EGD and identify BE in patients who do not report chronic GERD symptoms yet have three or more non-GERD risk factors that place them at risk for developing adenocarcinoma. The majority of BE that is identified by this approach is short segment BE. Full scale multi-center studies in an adult population with additional risk factors can help more precisely determine the predictive values and develop non-endoscopic screening strategies for patients without chronic GERD symptoms who are nevertheless at risk for developing cancer.

## Data Availability

All data produced in the present work are contained in the manuscript

## Abbreviations

BE: Barrett’s Esophagus
EAC: Esophageal Adenocarcinoma
EGD: Esophagogastroduodenoscopy
HGD: High Grade Dysplasia
LGD: Low Grade Dysplasia

## REFERENCES

1. Curtius K, Rubenstein JH, Chak A, Inadomi JM. Computational modelling suggests that Barrett’s oesophagus may be the precursor of all oesophageal adenocarcinomas. Gut. Nov 24 2020;70(8):1435–40. doi:gutjnl-2020-321598 [pii] 10.1136/gutjnl-2020-321598

2. Shaheen NJ, Falk GW, Iyer PG, Gerson LB, American College of G. ACG Clinical Guideline: Diagnosis and Management of Barrett’s Esophagus. Am J Gastroenterol. Jan 2016;111(1):30-50; quiz 51. doi:10.1038/ajg.2015.322

3. American Gastroenterological A, Spechler SJ, Sharma P, Souza RF, Inadomi JM, Shaheen NJ. American Gastroenterological Association medical position statement on the management of Barrett’s esophagus. Gastroenterology. Mar 2011;140(3):1084–91. doi:10.1053/j.gastro.2011.01.030 S0016-5085(11)00084-9 [pii]

4. Asge Standards Of Practice C, Qumseya B, Sultan S, et al. ASGE guideline on screening and surveillance of Barrett’s esophagus. Gastrointest Endosc. Sep 2019;90(3):335–359 e2. doi:S0016-5107(19)31704-3 [pii] 10.1016/j.gie.2019.05.012

5. Chak A, Faulx A, Eng C, et al. Gastroesophageal reflux symptoms in patients with adenocarcinoma of the esophagus or cardia. Cancer. Nov 1 2006;107(9):2160–6. doi:10.1002/cncr.22245

6. Lagergren J, Bergstrom R, Lindgren A, Nyren O. Symptomatic gastroesophageal reflux as a risk factor for esophageal adenocarcinoma. N Engl J Med. Mar 18 1999;340(11):825–31.

7. Moinova HR, LaFramboise T, Lutterbaugh JD, et al. Identifying DNA methylation biomarkers for non-endoscopic detection of Barrett’s esophagus. Science translational medicine. Jan 17 2018;10(424)doi:10.1126/scitranslmed.aao5848

8. Moinova HR, Verma S, Dumot J, et al. Multicenter, Prospective Trial of Nonendoscopic Biomarker-Driven Detection of Barrett’s Esophagus and Esophageal Adenocarcinoma. Am J Gastroenterol. Nov 1 2024;119(11):2206–2214. doi:10.14309/ajg.000000000000285000000434-990000000-01147 [pii]

9. Greer KB, Blum AE, Faulx AL, et al. Nonendoscopic Screening for Barrett’s Esophagus and Esophageal Adenocarcinoma in At-Risk Veterans. Am J Gastroenterol. Mar 1 2025;120(3):545–553. doi:10.14309/ajg.000000000000296200000434-990000000-01238 [pii]

10. Locke GR, Talley NJ, Weaver AL, Zinsmeister AR. A new questionnaire for gastroesophageal reflux disease. Mayo Clin Proc. Jun 1994;69(6):539–47. doi:S0025-6196(12)62245-9 [pii] 10.1016/s0025-6196(12)62245-9

11. Sharma P, Dent J, Armstrong D, et al. The development and validation of an endoscopic grading system for Barrett’s esophagus: the Prague C & M criteria. Gastroenterology. Nov 2006;131(5):1392–9. doi:S0016-5085(06)01791-4 [pii] 10.1053/j.gastro.2006.08.032

12. Rubenstein JH, Morgenstern H, Appelman H, et al. Prediction of Barrett’s esophagus among men. Am J Gastroenterol. Mar 2013;108(3):353–62. doi:10.1038/ajg.2012.446ajg2012446 [pii]

13. Asge Standards Of Practice C, Qumseya B, Sultan S, et al. ASGE guideline on screening and surveillance of Barrett’s esophagus. Gastrointest Endosc. Sep 2019;90(3):335–359 e2. doi:S0016-5107(19)31704-3 [pii] 10.1016/j.gie.2019.05.012

14. Muthusamy VR, Wani S, Gyawali CP, Komanduri S, Participants CBsECC. AGA Clinical Practice Update on New Technology and Innovation for Surveillance and Screening in Barrett’s Esophagus: Expert Review. Clin Gastroenterol Hepatol. Dec 2022;20(12):2696–2706 e1. doi:10.1016/j.cgh.2022.06.003

15. Chandar AK, Keerthy K, Gupta R, et al. Patients with Esophageal Adenocarcinoma with prior GERD symptoms are similar to those without GERD: A Cross-sectional Study. Am J Gastroenterol. Nov 17 2023;doi:10.14309/ajg.000000000000259300000434-990000000-00945 [pii]

16. Chandar AK, Low EE, Singer ME, Yadlapati R, Singh S. Estimated Burden of Screening for Barrett’s Esophagus in the United States. Gastroenterology. Mar 30 2023;doi:S0016-5085(23)00537-1 [pii] 10.1053/j.gastro.2023.03.223

17. Buas MF, Vaughan TL. Epidemiology and risk factors for gastroesophageal junction tumors: understanding the rising incidence of this disease. Semin Radiat Oncol. Jan 2013;23(1):3–9. doi:S1053-4296(12)00088-4 [pii] 10.1016/j.semradonc.2012.09.008

